# Multi-ancestry Genome-wide Association Study of Creatine Kinase Highlights the Genetic Basis of Muscle Damage

**DOI:** 10.1101/2025.09.24.25335552

**Authors:** Gang Chen, Sizheng S. Zhao, Hector Chinoy, James B Lilleker, Weijie Liu, Yuhui Li, Andrew P. Morris, Janine A. Lamb

## Abstract

Serum creatine kinase (CK) is a routinely measured biomarker of muscle damage, yet the genetic factors underlying inter-individual variation in CK levels remain poorly defined. Here we present the largest multi-ancestry genome-wide association meta-analysis of serum CK to date, comprising 237,255 participants spanning Admixed American, African American, East Asian, European and Middle Eastern populations. We identify 107 independent loci at genome-wide significance (P<5x10^-8^), 98 of which are previously unreported, with pronounced enrichment for genes expressed in skeletal and cardiac muscle and overlap with pathways related to muscle structure and function. Notably, eight loci map to genes implicated in Mendelian myopathies, underscoring a continuum from common regulatory variation to rare pathogenic mutations. Integrative quantitative trait locus (QTL)-based Mendelian randomization and colocalization implicate several genes in CK regulation, most prominently *SMAD3*, *KLF5* and *STAT3* within the transforming growth factor beta signalling pathway. CK levels show positive genetic correlations with traits reflecting tissue damage and muscle mass, and negative correlations with C-reactive protein, indicating pleiotropic effects on muscle biology and enzyme clearance. Together, these findings delineate the genetic architecture of serum CK across diverse populations and provide insight into the genetic basis of muscle-damage risk and subclinical CK elevation.

## Introduction

Creatine kinase (CK) is a key enzyme that catalyses the adenosine triphosphate (ATP)-dependent phosphorylation of creatine, thereby sustaining energy homeostasis in tissues with high and fluctuating metabolic demands, notably skeletal muscle, myocardium and brain^1^. Elevated serum CK is a well-established biomarker of tissue damage, observed in conditions ranging from benign myalgia and statin-associated myopathy to severe rhabdomyolysis^1,2^. Accordingly, CK measurement is routinely employed in the clinical assessment of muscle pathology. However, serum CK concentrations exhibit substantial inter-individual variability, influenced by factors such as age, sex, genetic ancestry, physical activity, and muscle mass^3^. Baseline levels are typically higher in males than in females, and individuals of Black African ancestry often present with markedly elevated CK relative to other ancestry groups^3^. This heterogeneity complicates the interpretation of CK as a diagnostic marker, as physiological elevations may be difficult to distinguish from pathological increases.

Genome-wide association studies (GWAS) in individuals of European ancestry have identified multiple genetic loci associated with serum CK levels, including *ANO5*, *CD163*, and *LILRB5*^4,5^. The largest study to date, comprising 63,159 Icelandic individuals, incorporated stratification by statin use to investigate CK variation^4^. One of the strongest associations was observed for a common missense variant in *LILRB5*. Further studies implicated this *LILRB5* variant in increased susceptibility to statin intolerance and myalgia^6^. Variants in *ANO5*, a gene associated with adult-onset muscular dystrophies^7^, also demonstrated modest associations with elevated CK levels, suggesting a continuum of genetic influences spanning subclinical biochemical alterations to myopathic disease. These findings reinforce the utility of CK as a sensitive biomarker of subclinical muscle damage and point to its potential in revealing the molecular basis of mild, often unrecognized, muscle pathologies in the general population.

Large-scale biobanks have catalysed major advances in GWAS of circulating biomarkers by integrating genomic data with detailed clinical and phenotypic information. Analyses of circulating biomarkers, including C-reactive protein (CRP) and troponin, in cohorts such as UK Biobank and INTERVAL have identified genetic loci linked to inflammation and cardiac injury, providing insights into disease pathophysiology^8,9^. More recent GWAS in extensively phenotyped cohorts—such as the Million Veteran Program (MVP) and BioBank Japan (BBJ) —have leveraged electronic health records (EHRs) and standardized laboratory assays to map loci influencing serum CK levels across diverse ancestries^10,11^. Despite these advances, most studies have prioritized statistical associations over functional annotation or mechanistic interpretation. Furthermore, reliance on single biobank datasets limits the resolution of causal inference and reduces the generalizability of findings across populations.

To address these gaps, we conducted the largest multi-ancestry GWAS meta-analysis of serum CK levels to date, including 237,255 individuals from diverse populations. Our analyses aimed to define the genetic architecture of CK, uncover biological pathways underlying its regulation, and clarify its relationship to muscle damage susceptibility. These findings provide a framework for understanding CK as a biomarker of muscle damage and its broader clinical implications.

## Results

### Analytical Framework and Cohort Concordance

We curated GWAS summary statistics for serum CK levels from six ancestry-specific cohorts from four biobanks: MVP^11^ (Admixed American, African American, and European ancestry), BBJ^10^ (East Asian ancestry), Vanderbilt University Medical Centre^12^ (VUMC; European ancestry), and Qatar Genome Program^13^ (QGP; Middle Eastern ancestry), comprising a total of 237,255 individuals (Supplementary Table 1). To identify genetic loci associated with serum CK and explore their functional relevance, we implemented a multi-step analytical framework integrating GWAS meta-analysis, functional annotation, enrichment analyses for Mendelian muscle disease genes, candidate gene prioritization, and genetic correlation (Figure 1).

**Figure 1.**
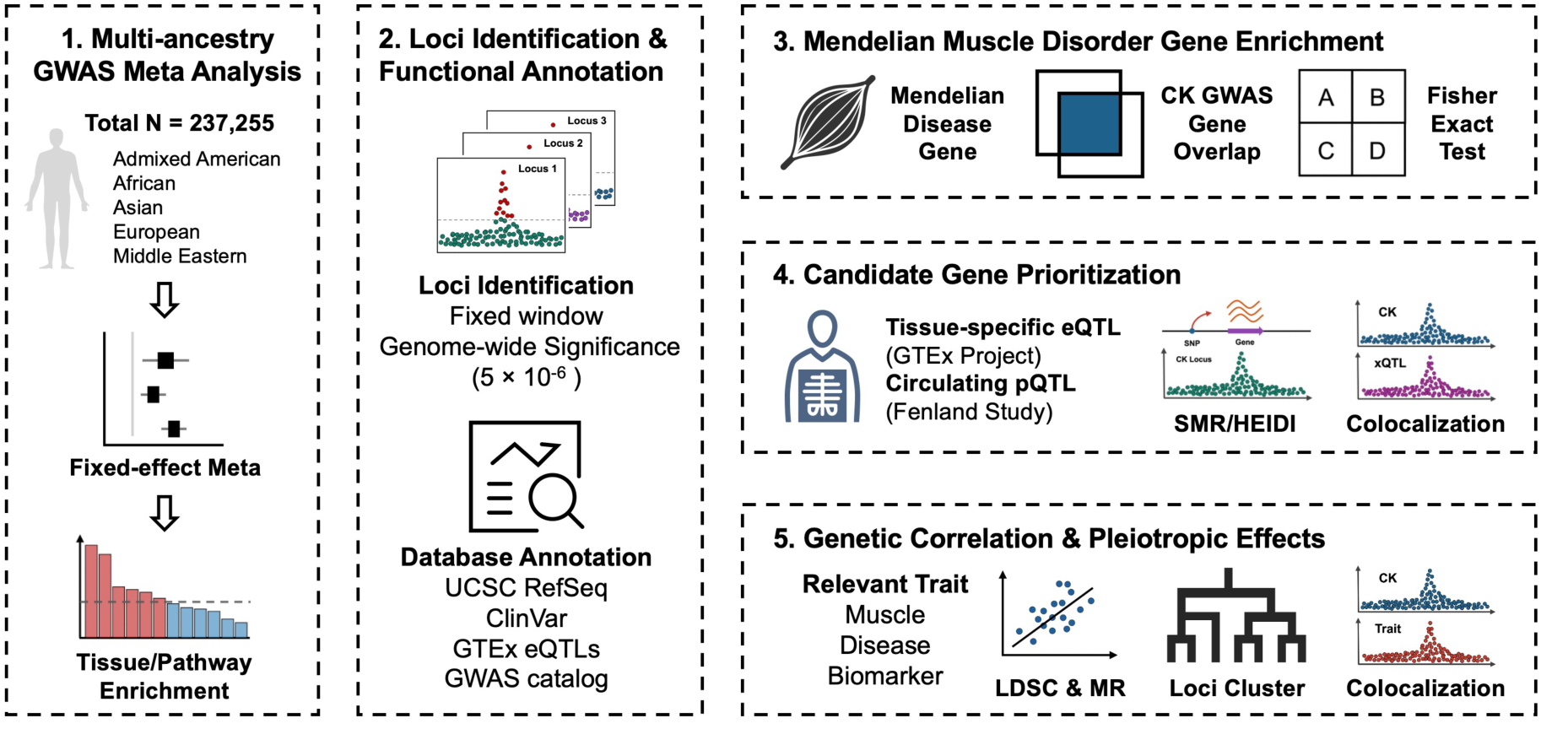
Overview of multi-ancestry GWAS and functional analyses of serum CK associated loci. We performed a fixed-effects meta-analysis of GWAS summary statistics for CK levels from six cohorts across diverse ancestries. Associated loci were annotated and tested for enrichment in Mendelian muscle disorder genes. Candidate genes were prioritized using eQTL and pQTL data. Shared genetic architecture with related traits was explored through LD score regression, Mendelian randomization, and colocalization. Loci were further clustered according to pleiotropic profiles.

Standardized quality control procedures were applied uniformly across all datasets (Methods; Supplementary Figures 1, 2). To assess the consistency of CK associations across cohorts, we compared effect sizes for independent lead variants and observed moderate to high correlations across most ancestry groups (Supplementary Figure 3). The highest pairwise concordance was observed between the European ancestry subset of MVP and VUMC (Pearson’s correlation coefficient (*r*) = 0.859, *P* = 2.94 × 10^-20^), whereas the lowest concordance was noted between VUMC and the African American ancestry subset of MVP cohort (*r* = 0.141, *P* = 0.297).

### Multi-Ancestry Meta-analysis and Significant Muscle Enrichment

We conducted the largest multi-ancestry GWAS meta-analysis of serum CK to date, leveraging cross-population linkage disequilibrium (LD) to improve fine-mapping and statistical power. Using Stouffer’s method to account for different trait transformations across biobanks, the primary meta-analysis encompassed 17,355,739 single nucleotide polymorphisms (SNPs) across 237,255 individuals and showed modest genomic inflation (λ = 1.13; Supplementary Table 2). The Manhattan plot of the multi-ancestry meta-analysis is shown in Figure 2A. Additional multi-ancestry meta-analyses of biobanks using the same trait transformation (i.e. excluding BBJ), as well as a European ancestry-specific meta-analysis, were performed using an inverse-variance weighted (IVW) approach (Supplementary Table 2 and Supplementary Figure 4). SNP-based heritability estimates from LD Score Regression (LDSC) were similar across European ancestry (VUMC and MVP European) and East Asian ancestry (BBJ) cohorts, at approximately 0.10 (Supplementary Table 3).

**Figure 2.**
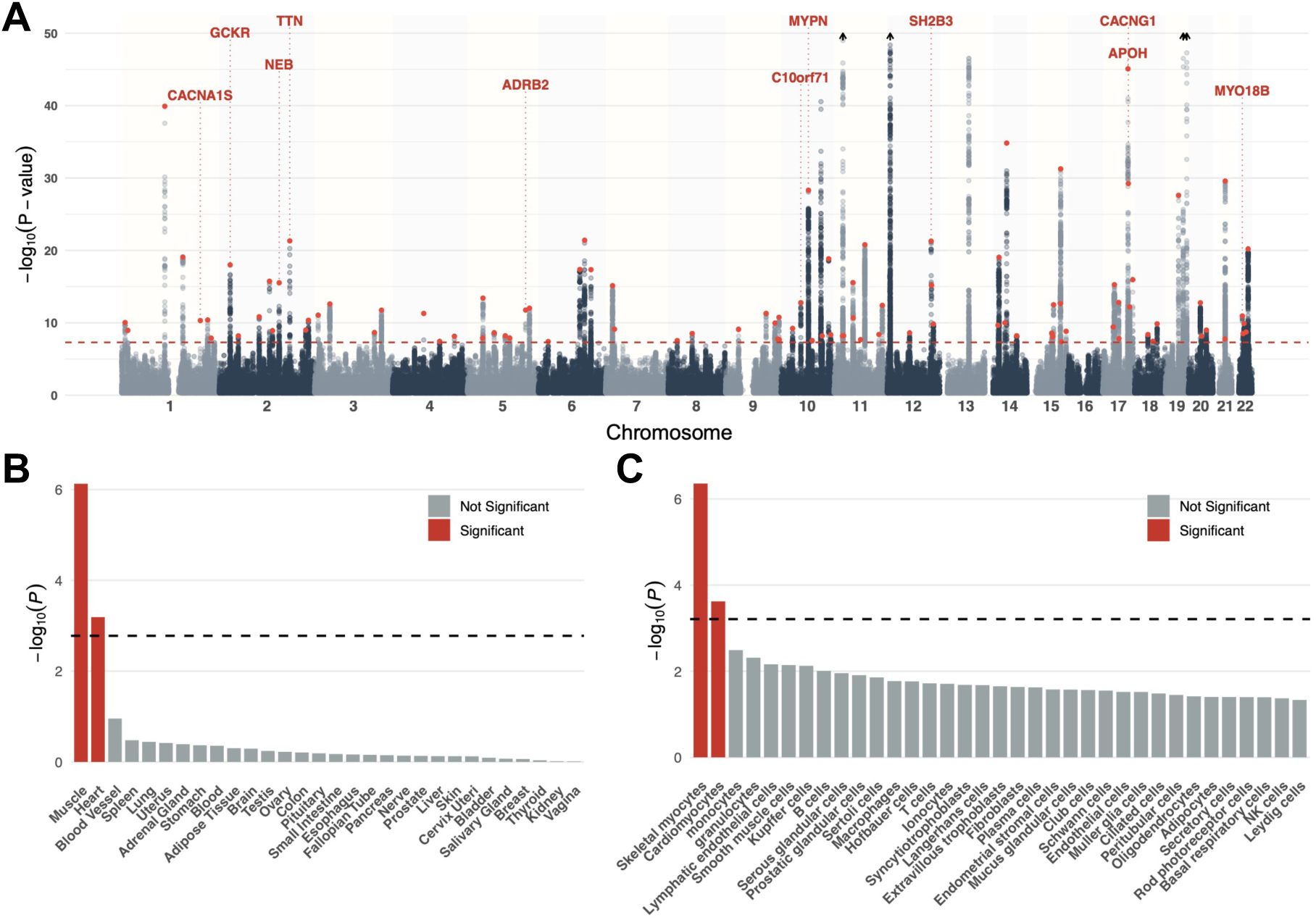
Multi-ancestry genome-wide association and tissue/cell-type enrichment analyses. (A) Manhattan plot of multi-ancestry GWAS results for CK across all autosomes; the red dashed line denotes the genome-wide significance threshold (P = 5 × 10^−8^). Novel lead SNPs are shown in red; for clarity only novel loci with exonic lead SNPs are annotated with mapped genes. Four known loci exceeding −log_10_(P) > 50 are indicated with arrows. (B & C) MAGMA tissue and cell-type enrichment analysis based on (B) GTEx expression data, (C) Human Protein Atlas single-cell expression profiles; the dashed line indicates the Bonferroni-corrected significance threshold, and significant tissues and cell types are shown in red.

Due to differences in LD structure, downstream functional analyses were based on the European ancestry-specific meta-analysis, which accounted for 38% of the total sample size. Gene- and gene-set enrichment analyses were conducted using Multi-marker Analysis of GenoMic Annotation (MAGMA), mapping SNP associations to 18,649 protein-coding genes. Ninety-nine genes surpassed the Bonferroni-corrected significance threshold (*P* ≤ 2.69 × 10^-6^; Supplementary Table 4), with *CKM* (*P* = 5.55 × 10^-17^), *WDR73* (*P* = 1.61 × 10^-15^), and *LILRB2* (*P* = 3.94 × 10^-15^) among the top associations. Tissue-level expression analysis using Genotype-Tissue Expression (GTEx) project data revealed significant enrichment in skeletal muscle (*P* = 7.44 × 10^-7^) and heart atrial appendage (*P* = 6.46 × 10^-4^) (Figure 2B; Supplementary Table 5). Single-cell transcriptomic data from the Human Protein Atlas further supported these findings, showing significant enrichment in skeletal myocytes ( *P* = 4.39 × 10^-7^) and cardiomyocytes ( *P* = 2.39 × 10^-4^) among 81 evaluated cell types, with monocytes ranking third (*P* = 3.23 × 10^-3^), though not significant after multiple testing correction (Figure 2B; Supplementary Table 6). Pathway enrichment analysis highlighted Gene Ontology biological processes related to the negative regulation of muscle adaptation (*P* = 1.05 × 10^-6^), negative regulation of cardiac muscle adaptation (*P* = 1.59 × 10^-6^), and striated muscle cell development (*P* = 2.43 × 10^-6^) as the most significantly enriched (Supplementary Table 7).

### Functional Landscape of CK-Associated Genetic Loci

In the multi-ancestry GWAS meta-analysis comprising all cohorts, we identified 107 independent loci at genome-wide significance (*P* < 5 × 10^-8^), defined as regions within ±500 kilobases (kb) of each lead variant. Of these, 98 loci represent novel associations not previously reported in earlier serum CK GWAS^4^,^5^,^14^. To assess their functional relevance, we annotated the 107 lead SNPs using publicly available resources (Supplementary Table 8), which informed downstream analyses.

Among the 107 loci, 15 lead variants mapped to exonic regions. The CK signals previously reported in *CKM* (*P* = 1.22 × 10^-219^), *LILRB5* (*P* = 1.11 × 10^-183^), and *ANO5* (*P* = 5.79 × 10^-60^) were replicated with the strongest associations in the multi-ancestry meta-analysis. All 15 exonic lead variants were missense, and ClinVar annotations indicate that 11 have previously been linked to Mendelian disorders, although mostly classified as benign. Notably, the newly identified rs3850625 (*P* = 4.89 × 10^-11^, p.R1539C) in *CACNA1S* has been implicated in congenital myopathy, malignant hyperthermia susceptibility, and periodic paralysis. Additional conditions associated with exonic variants include *MYPN*-related myopathy and dilated cardiomyopathy (for example, rs10997975 in *MYPN p.S691N*, *P* = 4.86 × 10^-29^), as well as disorders involving *TTN* (rs12463674, p.I17160T, *P* = 4.78 × 10^-22^), *MYO18B* (rs133885, p.G44E, *P* = 1.15 × 10^-11^), *NEB* (rs13013209, p.K2613N, *P* = 2.98 × 10^-16^) and *ANO5* (rs7481951, p.L321F, *P* = 5.79 × 10^-60^). Furthermore, an exonic variant in the non-coding RNA gene *TARID*, rs2277083 (*P* = 4.51 × 10^-18^), having prior ClinVar submissions referencing dilated cardiomyopathy 1J, was among the loci detected.

The majority of association signals arose from intronic regions, including 48 novel loci (Supplementary Table 8). Several intronic lead variants were coincidential with muscle expression quantitative trait loci (eQTLs), implicating genes such as *STAT3* (rs3736161, *P* = 1.54 × 10^-13^), *SMAD3* (rs12901499, *P* = 3.20 × 10^-13^) and *CSF1* (rs333947, *P* = 1.22 × 10^-40^). Intergenic loci also occurred in the same region as muscle eQTL associations; for example, rs11714943 ( *P* = 1.83 × 10^-12^), located between *PRKCI* and *SKIL*, were coincidental with an eQTL for *SKIL*. Similarly, the previously reported intronic variant rs9543398 (*P* = 2.91 × 10^-47^) in LINC00392 was coincidental with a muscle eQTL for *KLF5*.

Annotation against the GWAS Catalog revealed extensive pleiotropy among loci associated with serum CK levels. These loci are frequently linked to traits related to tissue damage, lipid metabolism, body composition, and cardiovascular phenotypes. Exonic variants in genes with muscle-specific expression displayed particularly strong pleiotropic effects. For instance, rs12975366 (*P* = 1.11 × 10^-183^) in *LILRB5* is associated with high-density lipoprotein (HDL) cholesterol and leukocyte immunoglobulin-like receptor subfamily B member 5 levels, whereas rs10997975 in *MYPN* associates with hip circumference (adjusted for body mass index), height and serum creatinine concentration. Similarly, rs3850625 in *CACNA1S* shows associations with aspartate aminotransferase (AST) levels, serum creatinine and whole-body fat-free mass.

### Shared Genetic Architecture with Mendelian Muscle Disorders

ClinVar annotations indicated a potential link between loci associated with serum CK levels and genes implicated in Mendelian muscle disorders. To investigate this relationship, we performed an enrichment analysis using a curated set of muscle-related Mendelian disorder genes from the Online Mendelian Inheritance in Man (OMIM) database (Supplementary Table 9). Among the 173 genes in this set, 8 overlapped with the 68 genes mapped to lead variants identified in the multi-ancestry CK GWAS meta-analysis (Table 1). This overlap represented a significant enrichment of CK-associated genes among those implicated in Mendelian muscle disorders (Fisher’s exact test, odds ratio [OR] = 13.99, 95% confidence interval [CI] = 5.68–29.96, *P* = 3.38 × 10^-7^). Five of the CK-associated loci contained nonsynonymous lead variants; all five are reported in ClinVar for relevant Mendelian conditions. In contrast, the three intronic lead variants lacked corresponding ClinVar annotations.

To further validate these findings, we repeated the enrichment analysis using all significant GWAS variants, which mapped to 221 CK-associated genes. Of these, 15 overlapped with the Mendelian muscle gene set, again indicating significant enrichment (Fisher’s exact test, OR = 7.91, 95% CI = 4.25–13.75, *P* = 5.18 × 10^-9^). The full list of genes and associated disorders is provided in Supplementary Table 10.

**Table 1.** Lead SNPs at CK-associated loci and their mapped genes with Mendelian muscle disease associations. SNP: lead variant representing the CK-associated locus; Chr: chromosome; Pos: genomic position (GRCh37, base pairs); Ref/Alt: reference and alternate alleles; Freq: frequency of the alternate allele in the multi-ancestry meta-analysis; P: association P-value for CK; Func: functional annotation of the lead SNP; Gene: gene mapped to the lead SNP; OMIM Disease: Mendelian muscle disease(s) associated with the gene in the OMIM database

| SNP | Chr | Pos | Ref | Alt | Freq | P | Func | Gene | OMIM Disease |
| --- | --- | --- | --- | --- | --- | --- | --- | --- | --- |
| **rs3850625** | 1 | 201,016,296 | G | A | 0.07 | $4.89 \times 10^{-11}$ | exonic | CACNA1S | Malignant hyperthermia susceptibility 5; Congenital myopathy 18 due to dihydropyridine receptor defect |
| **rs13013209** | 2 | 152,500,449 | C | G | 0.33 | $2.98 \times 10^{-16}$ | exonic | NEB | Nemaline myopathy 2, autosomal recessive |
| **rs12463674** | 2 | 179,432,185 | A | G | 0.16 | $4.78 \times 10^{-22}$ | exonic | TTN | Cardiomyopathy, dilated, 1G; Congenital myopathy 5 with cardiomyopathy; Cardiomyopathy, familial hypertrophic, 9; Muscular dystrophy, limb-girdle, autosomal recessive 10; Myopathy, myofibrillar, 9, with early respiratory failure |
| **rs237870** | 3 | 8,779,066 | T | C | 0.28 | $8.84 \times 10^{-12}$ | intronic | CAV3 | Myopathy, distal, Tateyama type; Cardiomyopathy, familial hypertrophic |
| **rs10997975** | 10 | 69,933,921 | G | A | 0.40 | $4.86 \times 10^{-29}$ | exonic | MYPN | Cardiomyopathy, familial restrictive, 4; Cardiomyopathy, hypertrophic, 22; Cardiomyopathy, dilated, 1KK; Congenital myopathy 24 |
| **rs72842207** | 10 | 121,433,675 | C | T | 0.17 | $1.43 \times 10^{-19}$ | intronic | BAG3 | Cardiomyopathy, dilated, 1HH; Myopathy, myofibrillar, 6 |
| **rs7481951** | 11 | 22,271,870 | A | T | 0.34 | $5.79 \times 10^{-60}$ | exonic | ANO5 | Muscular dystrophy, limb-girdle, autosomal recessive 12; Miyoshi muscular dystrophy 3 |
| **rs11081948** | 18 | 33,987,478 | A | G | 0.42 | $4.19 \times 10^{-9}$ | intronic | FHOD3 | Cardiomyopathy, familial hypertrophic, 28 |

### Mendelian Randomization and Colocalization Prioritize Candidate Genes

Functional annotation highlighted that multiple CK lead SNPs were coincidental with eQTLs across various tissues, particularly in muscle. We performed molecular QTL based Mendelian randomization (SMR) and colocalization analyses, which further highlighted candidate genes potentially involved in muscle damage and elevated CK levels. The heat map in Figure 3A summarizes the quantitative trait loci (QTL)-based SMR and colocalization results. After correction for multiple testing (*P_SMR_* < 1.70 × 10^-6^), we identified 42 significant SMR associations, predominantly in skeletal muscle, whole blood and subcutaneous adipose tissue. Of these, 23 associations passed the heterogeneity in dependent instruments (HEIDI) test (*P_HEIDI_* > 0.05), indicating consistency with vertical pleiotropy (Supplementary Table 11). Notably, the *LILRB5* protein quantitative trait locus (pQTL) emerged as the most significant association (*P_SMR_* = 3.11 × 10^-137^, *β_SMR_* = 0.104, *SE_SMR_* = 0.004). Additionally, *KLF5* (*P_SMR_* = 5.23 × 10^-19^, *β_SMR_* = −0.155, *SE_SMR_* = 0.017), *SMAD3* (*P_SMR_* = 1.14 × 10^-6^, *β_SMR_* = −0.108, *SE_SMR_* = 0.022) and *ALPK3* (*P_SMR_* = 1.81 × 10^-13^, *β_SMR_* = −0.160, *SE_SMR_* = 0.022) appeared exclusively in muscle-specific SMR analyses. Several genes were implicated across multiple tissues, including *ANO5*, which showed consistent associations in whole blood, liver, heart and adipose tissue, but not in skeletal muscle. The expression of *C1QTNF4* was associated with CK in all seven tissues analysed.

**Figure 3.**
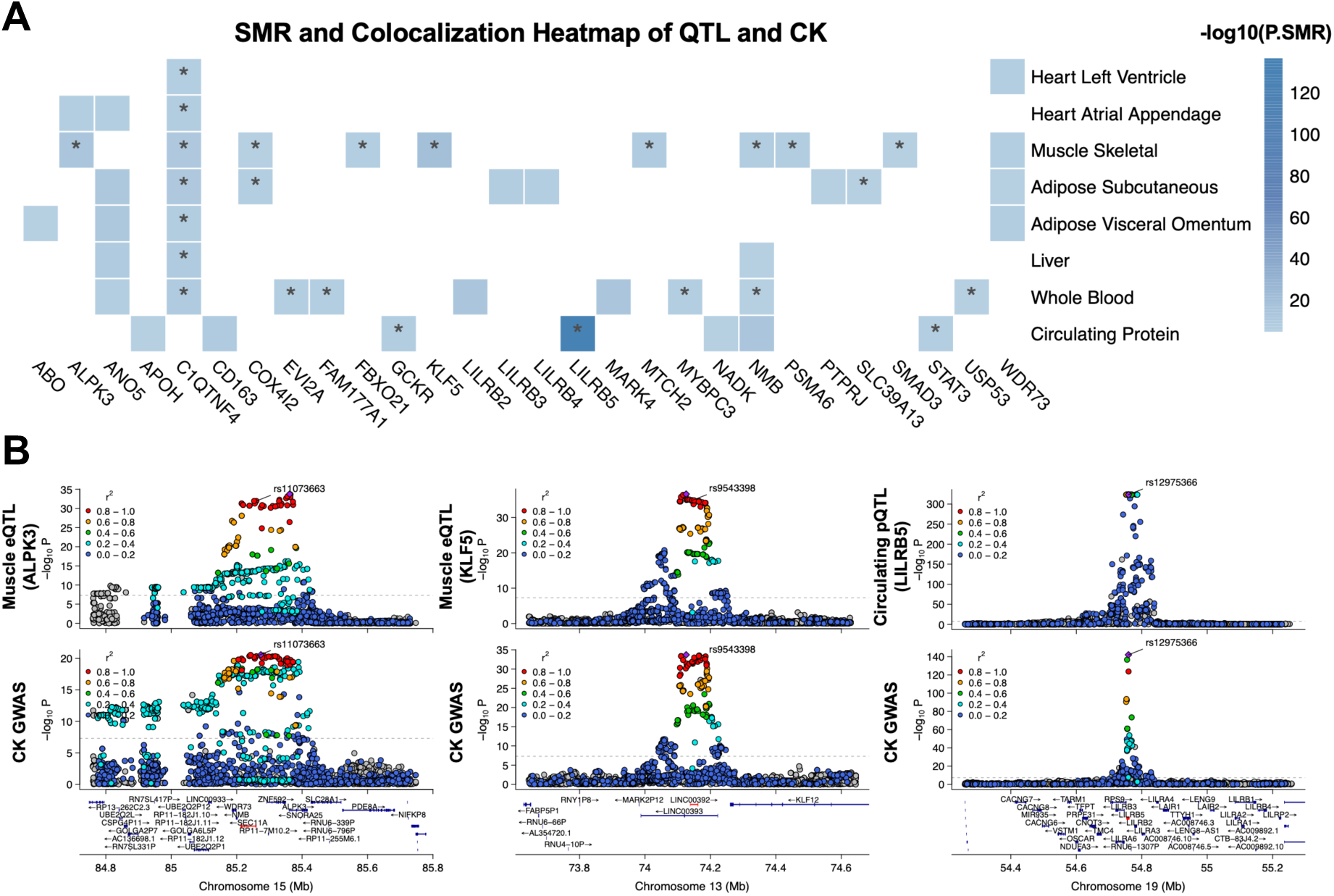
Mendelian Randomization and colocalization analyses implicate candidate genes for serum CK. (A) Heatmap of SMR associations (−log_10_P) between eQTLs and pQTLs across heart, skeletal muscle, adipose, liver, whole blood, and circulating proteins versus CK GWAS signals. Asterisks indicate significant colocalization. (B) Regional plots for ALPK3 in muscle (rs11073663), KLF5 in muscle (rs9543398), and LILRB5 in plasma proteins (rs12975366), showing QTL signals above and CK GWAS signals below. Variants are colored by LD r^2^ with the lead SNP.

Colocalization analyses provided further support for the SMR findings, particularly in skeletal muscle (Figure 3A and Supplementary Table 12). Genes such as *ALPK3* (posterior probability for colocalization [PP_H4_] = 0.996), *SMAD3* (PP_H4_ = 0.917) and *KLF5* (PP_H4_ = 0.987) demonstrated high posterior probabilities for a shared causal variant influencing both gene expression in muscle and CK levels. Conversely, *LILRB2* exhibited a strong SMR signal in whole blood (*P_SMR_* = 9.86 × 10^-14^, *β_SMR_* = 0.473, *SE_SMR_* = 0.064) but lacked colocalization support (PP_H4_ = 1.92 × 10^-4^). The circulating GCKR (PP_H4_ = 0.977) and LILRB5 (PP_H4_ = 1.000) pQTLs were further supported by strong colocalization evidence (Supplementary Table 12). Representative colocalization plots for the eQTL of *ALPK3* and *KLF5* in muscle, as well as the pQTL of LILRB5, are shown in Figure 3B.

### Pleiotropic EOects of CK-Associated Genetic Loci

GWAS Catalog annotations indicated extensive pleiotropy among CK-associated loci, spanning biomarkers, hematological traits, muscle phenotypes, and disease outcomes, partially mirroring genome-wide genetic correlations. On this basis, we assembled a set of CK-related traits, including quantitative biomarkers, muscle phenotypes, and diseases for correlation and pleiotropy analyses.

Genetic correlations between serum CK levels and related traits were evaluated using LDSC in the European ancestry cohorts (Figure 4A). Of the 42 traits assessed, nine showed significant genetic correlations after Bonferroni correction (*P* < 1.19 × 10^-3^). The strongest positive correlation was observed with AST levels (*r*_g_ (*LDSC*) = 0.410, *P* = 6.88 × 10^-15^), followed by muscle-related traits, including hand grip strength (*r*_g_ = 0.187, *P* = 2.35 × 10^-10^), arm fat-free mass (*r*_g_ = 0.144, *P* = 2.87 × 10^-7^), and basal metabolic rate (*r*g = 0.106, *P* = 8.14 × 10^-5^). CK levels were also significantly correlated with multiple circulating biomarkers, including alanine aminotransferase (ALT; *r*_g_ = 0.198, *P* = 3.60 × 10^-6^), creatinine (*r*_g_= 0.195, *P* = 3.87 × 10^-10^), albumin (*r*_g_= 0.189, *P* = 5.17 × 10^-6^), and cystatin C (*r*_g_ = −0.109, *P* = 8.00 × 10^-4^), as well as the inflammatory marker CRP (*r*_g_ = −0.185, *P* = 1.10 × 10^-8^). A full list of genetic correlations is provided in Supplementary Table 13.

To assess the directionality of these associations, we conducted two-sample Mendelian randomization (MR) analyses for traits used in the genetic correlation analysis (Supplementary Table 14). Significant causal effects (*P_IVW_* < 6.10 × 10^-4^) on CK were observed for AST (*P_IVW_* = 7.74 × 10^-9^, *β_IVW_* = 0.274), arm fat-free mass (*P_IVW_* = 5.15 × 10^-9^, *β_IVW_* = 0.170), hand grip strength (*P_IVW_* = 7.91 × 10^-5^, *β_IVW_* = 0.246), and creatinine (*P_IVW_* = 1.76 × 10^-4^, *β_IVW_* = 0.099). Conversely, when CK was used as the exposure, only AST showed significant evidence of a causal effect (*P_IVW_* = 1.17 × 10^-4^, *β_IVW_* = 0.164).

Instrumental variable plots and MR estimates for AST, hand grip strength, and arm fat-free mass are shown in Supplementary Figure 5. All traits demonstrated substantial heterogeneity across instrumental variables (Cochran’s Q test, *P* < 0.05), and Egger’s test indicated potential horizontal pleiotropy for AST both as exposure (*P_Egger_* = 0.001) and outcome (*P_Egger_* = 0.014).

**Figure 4.**
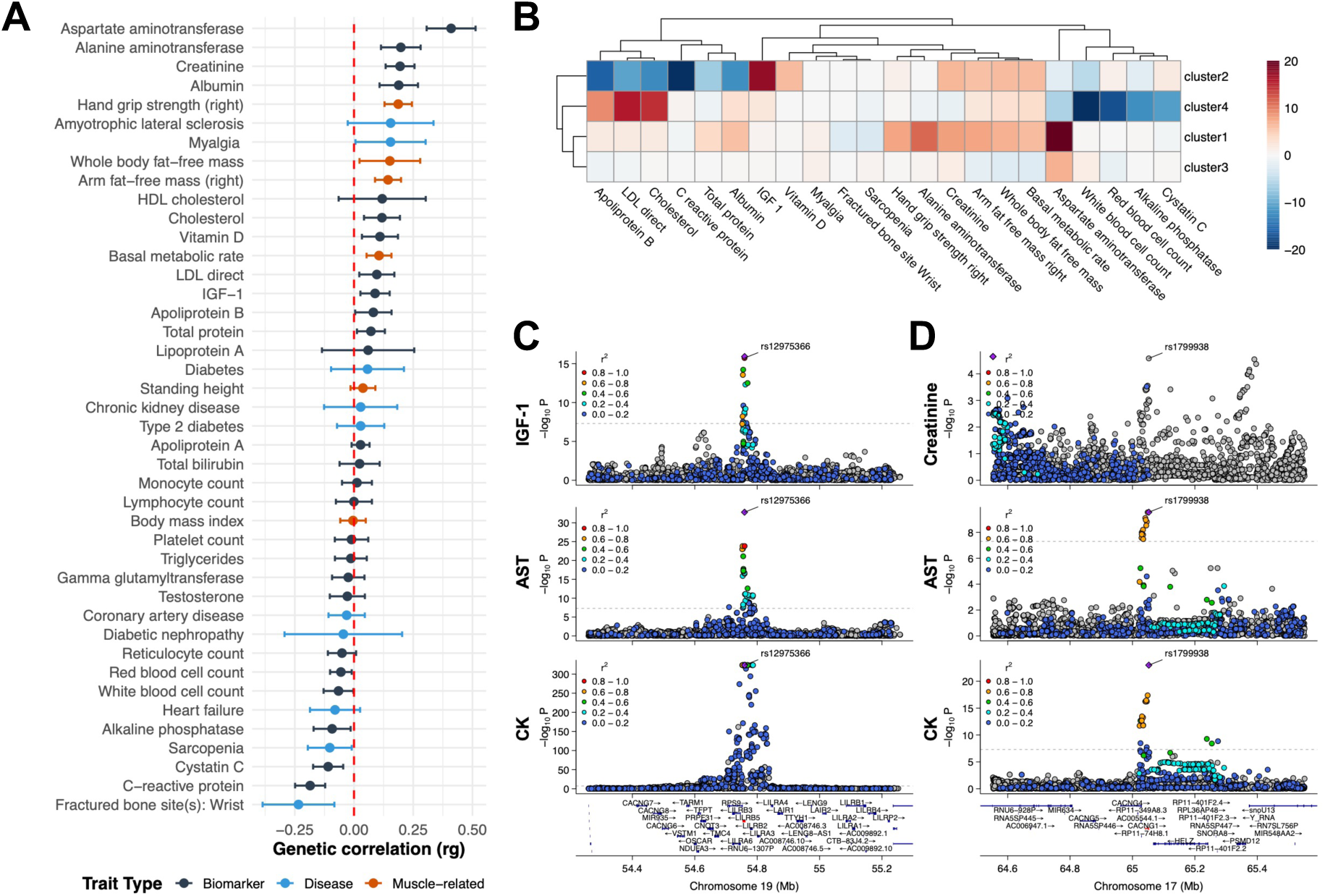
Genetic correlations and pleiotropic eHects of CK-associated loci. (A) Genome-wide genetic correlations between CK and 42 traits estimated by LDSC. Traits are grouped as biomarkers (black), diseases (blue), and muscle-related traits (red). Horizontal bars denote 95% CI; red dashed line, zero correlation. (B) Heat map of cluster–trait correlations for CK-associated loci; color indicates magnitude and direction. (C) Regional plot for rs12975366 showing colocalization of IGF-1, AST, and CK signals. Dotted line, genome-wide significance (P = 5 × 10^−8^); points colored by LD r^2^ with lead SNP. (D) Regional plot for rs1799938 demonstrating shared effects on creatinine, AST, and CK.

Effect sizes of lead SNPs across correlated traits further informed the functional characterization of CK-associated loci through clustering and multi-trait colocalization analyses. Eight lead SNPs lacking coverage across most traits and 20 traits with weak genetic correlations with CK (*P_LDSC_* > 0.05) were excluded. The remaining 99 lead variants were grouped into four clusters based on their effect sizes across 22 remaining traits (Supplementary Figure 6 and Supplementary Table 15). Notably, Clusters 1 and 3 accounted for the majority of lead variants (95 of 99). These two clusters showed consistent positive associations with AST (*P* = 8.76 × 10^−24^ for Cluster 1; *P* = 7.53 × 10^−8^ for Cluster 3), with Cluster 1 also exhibiting associations with ALT (*P* = 4.50 × 10^−12^) and creatinine (Figure 4B). By contrast, Clusters 2 and 4 exhibited negative correlations with

AST levels and white blood cell count (Figure 4B). Cluster 2 was also negatively associated with cholesterol and low-density lipoprotein (LDL) levels, whereas Cluster 4 showed positive correlations with these lipid traits. Cluster 2 includes rs1260326, a coding variant in *GCKR*. Cluster 4 comprises loci mapping to *SH2B3*, *PTPN11*, and *ABO*.

Multi-trait colocalization analysis confirmed shared genetic signals between CK and these traits (Supplementary Table 16), particularly AST and ALT. For example, the CK-associated locus rs7481951 in *ANO5* (posterior probability [PP] = 0.999) colocalized with both AST and ALT. The rs12975366 variant in *LILRB5* colocalized with AST and insulin-like growth factor 1 (IGF-1) (PP = 0.959), while rs1799938 in *CACNG1* colocalized with AST (PP = 0.835). Cross-trait regional plots of *LILRB5* and *CACNG1* are shown in Figure 4C & D. However, colocalization of CK signals in cluster 2 and 4 with other traits could not be confirmed.

## Discussion

Creatine kinase plays a central role in muscle energy metabolism and serves as a widely used clinical biomarker of muscle damage. Despite its clinical utility, serum total CK levels exhibit considerable inter-individual variability, limiting the ability to differentiate between normal physiological fluctuations and pathological elevations^3^. To elucidate the genetic architecture underlying CK variation, we performed the largest multi-ancestry GWAS meta-analysis of serum CK levels to date.

This study examines variation in serum CK levels within population biobanks, recognizing that genetic associations may differ in disease-specific contexts. Most samples were drawn from the BBJ and MVP cohorts. Although BBJ includes individuals with diverse clinical conditions, disease status was included as a covariate in CK GWAS, and the observed maximum CK value of 332 units per litre (U/L) suggests that extreme elevations associated with acute damage were likely absent. In MVP, individuals with markedly elevated CK, typically reflecting acute muscle damage, were excluded. These measures help to minimize confounding from disease-related elevations in CK. Additionally, a rank-based inverse normal transformation was applied in MVP, QGP, and VUMC cohorts to reduce the influence of outliers. These steps help ensure that the analysis reflects genetic influences on CK levels independent of severe muscle disease.

Ancestry-specific effect sizes of significant SNPs were moderately concordant in direction and magnitude across six cohorts and were combined in multi-ancestry meta-analysis. SNP-based heritability, estimated using LDSC, was consistent across the MVP European ancestry subset, VUMC and, BBJ, yielding an overall estimate of approximately 10%. MAGMA analyses revealed genome-wide enrichment across tissues, cell types, and pathways, with recurrent signals in muscle-related contexts. Pronounced muscle specificity was also observed for loci mapping to key muscle-function genes, including *ANO5*, *TTN*, and *MYPN*, with eQTL colocalization signals predominantly observed in muscle tissue compared to other tissues. Together, these findings suggest that the genetic regulation of serum CK levels in healthy individuals primarily reflects intrinsic muscle homeostasis. Although not statistically significant, enrichment in monocytes and granulocytes, ranking third and fourth, was comparable with previous GWAS findings in an Icelandic cohort and supports a potential role for monocyte/macrophage-mediated CK clearance^4^, in line with associations at *CSF1*.

We assessed genome-wide genetic correlations between serum CK and a broad range of physiological and disease-related traits. The strongest positive correlations were observed with liver enzymes, particularly AST, likely reflecting shared tissue origin due to muscle damage^15^. The colocalization of loci in muscle-specific genes, such as *CACNG1* and *ANO5*, suggests that the elevated AST levels originate from muscle release. Trait-based clustering further supported this link, with CK-associated loci generally aligning in effect direction with AST. In contrast, ALT showed a weaker correlation, likely due to its liver-specific expression^16^. CK also showed a positive genetic correlation with creatinine, a metabolite derived from muscle^17^. Among disease traits, nominal positive correlations were observed with amyotrophic lateral sclerosis and myalgia, though these did not reach statistical significance. Notably, CK was negatively correlated with wrist fracture, possibly reflecting underlying differences in muscle strength and reduced fracture risk^18^. This was supported by positive correlations between CK and both hand grip strength and fat-free mass, highlighting the influence of muscle mass and physical activity on serum CK levels^19^. CK was negatively correlated with inflammatory markers such as CRP and leukocyte count, showing agreement with reports linking lower CK levels to haematologic malignancies and increased immune-mediated enzyme clearance^20,21^. These findings imply that heightened immune activity may reduce CK levels through enhanced clearance mechanisms. Positive correlations were also observed with total cholesterol and LDL, suggesting an interaction between muscle and lipid metabolism. The significant correlation with albumin and cystatin C lack strong support in current literature and warrants further study using individual-level data. We note that these genome-wide correlations reflect average, intricate patterns of CK and related traits at the population level. However, locus-specific effects may vary in direction, as the two-sample MR analyses showed extensive instrumental variable heterogeneity and horizontal pleiotropy. The causality among these traits may need to be evaluated at the variant level and may differ across loci.

The multi-ancestry meta-analysis identified 107 loci associated with CK levels, including 98 not previously reported. Here, we focus on loci with strong prior links to muscle biology, exonic variants implicated in Mendelian disorders, and loci exhibiting distinct pleiotropic patterns across trait-defined clusters; additional loci are described in the Supplementary Note. Notably, multiple loci carrying exonic nonsynonymous variants were mapped to genes essential for muscle structure and sarcolemmal integrity. These CK loci genes were significantly enriched in a Mendelian muscle disorder gene set, which is consistent with those observed in other complex traits^22^. For example, the variant rs12463674 lies within a conserved A-band genomic region of the *TTN* gene, a known hotspot for titinopathy mutations^23^. Variants in *NEB* (rs13013209) and *MYPN* (rs10997975), core Z-line and sarcomere components, are associated with CK efflux and are annotated in ClinVar for benign myopathy^24,25^. A novel association at rs3850625 in *CACNA1S*, encoding the α1S subunit of the skeletal muscle L-type calcium channel, colocalizes between CK levels, grip strength, and lean mass, supporting a shared role in excitation–contraction coupling^26^. ClinVar links this variant to benign congenital myopathy and malignant hyperthermia susceptibility^26,27^. Similar associations were observed at rs1799938 in *CACNG1* and rs7481951 in *ANO5*, both involved in calcium homeostasis and membrane repair^28,29^. *ANO5* has been previously linked to muscular dystrophies and variation in CK levels^4,7^. Notably, additional signals were mapped to non-coding regions of ion channel genes, including *KCNMA1* and *KCNJ2*, further underscoring the importance of ion regulation. In addition to five OMIM-reported genes—*CACNA1S*, *MYPN*, *ANO5*, *NEB*, and *TTN*—numerous other loci were associated with skeletal and cardiac muscle phenotypes, although many remain functionally uncharacterized. An association was identified at rs133885 in *MYO18B*, involved in sarcomere assembly in fast skeletal muscle^30^; although associated with neurodevelopmental syndromes such as Klippel–Feil anomaly^31^, its role in muscle remains poorly defined. Lastly, rs10857472, a nonsynonymous variant in *C10orf71*, lies in a gene where frameshift mutations have been linked to dilated cardiomyopathy in humans, mice, and cardiac organoids^32^.

Although the identified exonic variants are common and likely not fully penetrant, their presence in genes linked to Mendelian myopathies suggests they may act as genetic modifiers or influence disease susceptibility under certain conditions. Variants in *TTN*, *NEB*, and *ANO5*—genes associated with recessive muscular disorders—could subtly disrupt sarcolemmal stability or calcium homeostasis^29,33^. These subclinical effects may increase vulnerability to muscle damage in response to stressors like intense exercise, infection, or medication. While the lack of individual-level and longitudinal data limits direct risk assessment, the involvement of these variants in structural and signaling pathways essential to muscle function supports a model in which common alleles influence muscle resilience. Elevated serum CK levels may serve as a biomarker of latent sarcolemmal fragility, unmasked by additional genetic or environmental factors. This model is consistent with clinical observations, where subclinical muscle disorders have emerged following exposure to agents such as statins. In one study of 110 individuals with drug-induced myopathies, 10% carried heterozygous or homozygous pathogenic mutations in genes known to cause these disorders^34^, highlighting the value of genetic testing even in mild or unexplained cases.

Candidate gene prioritization identified putative mediators of muscle damage within the genetic architecture of serum CK, particularly at non-coding loci. In addition to genes implicated in muscle function (*ALPK3*) and immune regulation (*LILRB5*), components of the transforming growth factor-beta (TGF-β) signalling pathway were recurrently highlighted, with several loci mapping to its regulatory genes. Among these, the CK signal at an intronic variant in *SMAD3* (rs12901499) colocalizes with its muscle eQTL, and eQTL-based MR supports a causal link between increased *SMAD3* expression and reduced CK levels. *SMAD3*, a central mediator of TGF-β signalling, is essential for muscle mass maintenance and satellite cell function^35^. Its loss leads to atrophy, impaired myoblast differentiation, and elevated myostatin expression, a known inhibitor of muscle growth^35,36^. Two additional loci further implicate TGF-β signalling. At *SKIL* (SnoN), muscle eQTLs were detected, although without colocalization with CK association signals. Given *SKIL*’s role as a SMAD-recruited transcriptional repressor^37^, and its degradation upon *SMAD3* activation, variation at this locus may modulate pathway output, pending functional validation. At *STAT3*, a key node in cytokine–TGF-β crosstalk^38^, colocalization and pQTL-MR suggest that genetically elevated circulating STAT3 is associated with lower CK levels. A locus near LINC00392, close to the 13q22.1 signal reported in an Icelandic CK study^4^, colocalizes with *KLF5*, whose increased expression in muscle is causally linked to reduced CK. *KLF5* is involved in exercise-induced lipid remodeling—promoting lipid oxidation acutely, and lipid synthesis with chronic training—and contributes to muscle atrophy through coordination with *FOXO1* and E3 ubiquitin ligases^39,40^. KLF5 also engages in bidirectional crosstalk with the TGF-β pathway: it promotes TGF-β expression and profibrotic signalling during fibroblast-driven remodeling, while active SMAD2/3– SMAD4 complexes recruit p300 to acetylate *KLF5*, forming a KLF5–SMAD–p300 complex that drives TGF-β target gene expression^41^. This positions *KLF5* as a potential integrator of TGF-β signalling with metabolic and proteolytic muscle responses, linking CK genetics to the broader TGF-β/SMAD axis involving *SMAD3*, *STAT3*, and *SKIL*. Other TGF-β–related genes, including *SMAD7* and *BACH1*, showed no eQTL colocalization, leaving their roles in CK regulation unresolved. Together, these findings highlight that these genetic variants may influence CK levels and potential muscle damage susceptibility though TGF-β signalling.

Analysis of pleiotropic effects revealed that most loci are primarily associated with relatively isolated muscle damage, as indicated by consistent effects on tissue damage biomarkers such as AST at the *CACNG1* and *LILRB5* loci. This pattern aligns with the established roles of genes at these loci in maintaining muscle structural integrity and contraction function. In contrast, four loci identified through trait-based clustering exhibited divergent pleiotropic signatures, indicative of more complex underlying biological processes. The variant rs1260326 in an exon of *GCKR* defines Cluster 2 and exhibits a concordant association with serum levels of CK and creatinine, but a discordant association with AST. This locus is also linked to systemic traits such as CRP and body mass index. *GCKR*, a key regulator of hepatic and pancreatic glucokinase, plays a central role in glucose metabolism and is a well-established locus for metabolic traits and disorders^42^. The genetic association with CK may reflect pleiotropic effects mediated through multiple metabolic pathways. Colocalization and the positive effect of circulating *GCKR* levels on CK suggest this locus may be a viable therapeutic target. The loci *SH2B3*, *PTPN11*, and *ABO* form Cluster 4, characterized by associations with both immune and metabolic traits. *SH2B3* is a negative regulator of cytokine signalling essential for haematopoiesis^43^, while *ABO* is best known for determining blood group. *PTPN11* encodes a tyrosine phosphatase involved in key signalling pathways and is associated with Noonan syndrome and acute myeloid leukaemia^44^. This cluster likely captures hematopoietic effects, consistent with observed reductions in CK in hematologic malignancies^20^. Outside of Cluster 4, the LINC02227/EBF1 locus also demonstrates haematopoietic relevance, showing colocalization with lymphocyte count. *EBF1* has been implicated in immune-related diseases such as acute lymphoblastic leukaemia^45^.

While our findings reflect the genetic architecture of population CK levels, we could not fully exclude individuals with acute conditions, and some residual influence from transient elevations may remain. Moreover, the lack of individual-level data limits deeper analyses of CK distributions, genotype-specific risk, and trait-adjusted models, which warrant future investigation.

In summary, we present the largest multi-ancestry meta-analysis of serum CK levels to date, identifying previously unreported genetic loci and providing preliminary insights into their potential roles in muscle damage. These results advance the clinical utility of CK as a biomarker and suggest its promise in genetic risk profiling for muscle-related disorders.

## Methods

### Cohorts, Phenotypes, and Genotyping

We performed a meta-analysis using six cohorts of ancestry-specific GWAS summary statistics from four biobanks: MVP^11^ (African, Admixed American, European ancestries), BBJ^10^ (East Asian ancestry), VUMC^12^ (European ancestry) and QGP^13^ (Middle Eastern ancestry), encompassing a total of 237,255 individuals. MVP is a large U.S.-based biobank with extensive phenotype data derived from EHRs^11^. BBJ provides deep clinical laboratory data and genome-wide genotyping^10^. VUMC is a tertiary medical centre in Nashville, Tennessee, with EHR-linked genomic data comprising diagnostic codes, laboratory results, and clinical narratives^12^. QGP is a population-based initiative conducting whole-genome sequencing with CK measured at Hamad General Hospital^13^. Genotyping in BBJ, MVP, and VUMC was performed using array-based platforms followed by imputation and quality control based on imputation accuracy.

Serum CK concentrations were measured in units of enzymatic activity per volume or U/L, derived from clinical laboratory assays or EHR data. In BBJ, CK values ranged from 7 to 332 U/L (median = 89 U/L); MVP excluded extreme outliers beyond six standard deviations. Trait normalization was performed using rank-based inverse normal transformation in QGP, MVP, and VUMC, and log-transformation followed by Z-score standardization in BBJ. Additional cohort-specific details are available in Supplementary Table 1.

### Quality Control and Data Harmonization

Quality control was conducted at both the cohort and meta-analysis levels using the EasyQC^46^ pipeline supplemented with custom scripts. At the cohort level, we harmonized input formats, excluded rare variants (minor allele frequency < 1%), structural variants, and SNPs with missing or non-ACGT alleles. SNP identifiers were standardized across cohorts, and all variants were mapped to the GRCh37 (hg19) reference genome. At the meta-analysis level, we evaluated data integrity and potential biases using a suite of diagnostic tools, including SE-N and P–Z plots, allele frequency concordance checks, and genomic inflation statistics.

### Lead Variant Concordance Across Cohorts

To evaluate consistency of genetic associations between cohorts, we compared effect estimates of lead variants across studies. Genome-wide significant variants (*P* < 5 × 10⁻⁸) were identified independently within each cohort and combined into a candidate lead variant list. To define independent signals, variants were clumped using LD estimates from the 1000 Genomes Project^47^ (all populations) with a 1 megabase (Mb) window and an R² threshold of 0.1. Pairwise concordance of effect directions and magnitudes was assessed by calculating Pearson’s correlation coefficients between effect sizes across cohorts.

### Meta-analysis of Genome-wide Association Statistics

We conducted meta-analyses using the fixed-effect IVW approach and the Stouffer’s method as implemented in METAL^48^. For the European ancestry meta-analysis, which included the MVP European ancestry and VUMC cohorts, we applied the IVW method. For the multi-ancestry meta-analysis, IVW was used when the BBJ cohort was excluded, whereas the Stouffer method was applied when BBJ was included, to account for differences in data transformation and effect size of BBJ. The largest multi-ancestry meta-analysis, including BBJ, was used for locus discovery, whereas the European-only meta-analysis was used for functional follow-up. The multi-ancestry analysis excluding BBJ provided complementary estimates of effect sizes. Variants located within the human leukocyte antigen region (chromosome 6: 25–34 Mb, hg19) were excluded from all analyses. All other variants were retained for subsequent locus discovery and functional annotation.

### Gene- and Gene-Set Enrichment Analysis

Gene- and gene-set association analyses were performed using MAGMA^49^ with summary statistics from the European GWAS meta-analysis. SNPs were mapped to protein-coding genes, and gene-level association statistics were calculated by aggregating SNP-wise effects while accounting for LD using the 1000 Genomes Project European reference panel, as implemented in Functional Mapping and Annotation (FUMA)^50^. Gene-level p-values were subsequently used to assess enrichment of predefined gene sets, including tissue- and cell type-specific expression profiles from the GTEx^51^ and the Human Protein Atlas^52^, as well as curated pathway sets available through FUMA^50^. Multiple testing correction for both gene- and gene-set analyses was applied using Bonferroni adjustment.

### Locus Discovery and Functional Annotation

Genome-wide significant loci were identified using the largest multi-ancestry meta-analysis dataset. For each chromosome, the variant with the smallest P value was designated as the lead SNP. A ±500 kb window was applied around each lead SNP, and all variants within this interval were assigned to the same locus. This procedure was repeated iteratively until no additional variants surpassed the genome-wide significance threshold (*P* < 5 × 10^−8^). To distinguish novel from previously reported associations, we curated a reference list of known variants from the prior CK-related GWAS^4,5,14^. Novel loci were defined as those without a previously reported association within ±1 Mb of the lead SNP.

Functional annotation of lead variants was performed using ANNOVAR^53^, mapping to genes in the UCSC refGene sequences (release 2020-08-22) and assessing pathogenicity information through ClinVar^54^ (version 2024-09-17). Previously reported associations were retrieved from the GWAS Catalog^55^, and regulatory effects were evaluated by integrating skeletal muscle eQTL from GTEx v8^51^.

### Enrichment in Muscle Mendelian Disease Gene Set

Genes associated with Mendelian disorders were obtained from the OMIM database^56^ and restricted to phenotypes with a confirmed molecular basis (phenotype mapping key = 3). The gene set was curated to include muscle-specific disorders by JBL. CK-associated loci were mapped to genes using UCSC refGene annotations, and intergenic loci were excluded. The primary analysis considered genes corresponding to lead variants, while a secondary analysis included all genome-wide significant variants (*P* < 5 × 10^−8^). Only protein-coding genes with an approved symbol from the HUGO Gene Nomenclature Committee were retained, defining a background of 17,554 genes. Enrichment of CK-associated genes within the Mendelian muscle disease gene set was evaluated using a 2 × 2 contingency table, and statistical significance was assessed with Fisher’s exact test (two-sided; *P* < 0.05).

### Gene Prioritization with Mendelian Randomization and Colocalization

We assessed causal relationships between gene expression and serum CK using SMR with the HEIDI test^57^ on European-ancestry GWAS summary statistics. Analyses were conducted with the SMR portal, which integrates pre-computed QTL data.

CK GWAS summary statistics were integrated with eQTL from GTEx v8 across relevant tissues (skeletal muscle, whole blood, heart atrial appendage, heart left ventricle, liver, visceral adipose, and subcutaneous adipose)^51^ and circulating pQTL from the Fenland study^58^. Associations were considered significant at *P* < 0.05 after Bonferroni correction; HEIDI *P* > 0.05 was interpreted as consistent with vertical pleiotropy rather than linkage.

Colocalization analyses were subsequently performed for 107 loci identified in the multi-ancestry meta-analysis (including BBJ) using the ‘coloc’ R package^59^. For each locus, colocalization was assessed within ±500 kb of the lead variant, comparing European-ancestry GWAS effects with GTEx eQTL data^51^ from corresponding tissues.

### Heritability and Genetic Correlation Estimation

SNP-based heritability was estimated separately in BBJ, VUMC, the MVP, and a European meta-analysis using LDSC^60^ with ancestry-matched pre-computed LD scores from the 1000 Genomes Project Phase 3 reference panels.

Genetic correlations between serum CK and related traits, including biomarkers, muscle phenotypes, and disease outcomes, were quantified using LDSC. The European summary statistics for these traits were curated from the GWAS Catalog^55^ and OpenGWAS^61^. To reduce bias from poorly imputed variants, analyses were restricted to high-quality variants across imputation panels, following LDSC best-practice recommendations^60^.

To further explore potential causal relationships, we conducted two-sample MR analyses^62^. Genome-wide significant variants associated with each exposure were selected as instruments and harmonized with outcome summary statistics. CK was assessed bidirectionally as both exposure and outcome. Causal estimates were evaluated using IVW as the primary method. Heterogeneity and directional pleiotropy were assessed using Cochran’s Q statistic and MR-Egger intercept^63^, respectively.

### Trait-based Clustering of CK-associated Loci

We clustered loci according to their effect sizes across traits related to CK. Effect estimates for CK were obtained from our European GWAS meta-analysis. Eight lead SNPs were excluded owing to limited coverage (reported < 30 traits out of 42 traits) and traits with weak genetic correlation to CK (*P_LDSC_* > 0.05) were also excluded. All effect sizes were aligned to the CK-increasing allele. For each locus–trait pair, we computed sample size adjusted z-scores as 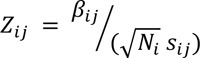, where *B_ij_* denotes the effect estimate, *s_ij_* its standard error, and *N_i_* the corresponding sample size. Missing values were imputed using the ‘ClustImpute’ R package^64^, which implements iterative k-means clustering while incorporating phenotypic correlations. The optimal number of clusters was determined with the ‘NbClust’ package^65^, based on consensus across 27 clustering indices. Associations between traits and cluster membership were assessed by linear regression. Heat maps were generated to visualize the clustering structure, with colour intensity reflecting both the direction of effect and the −log_10_-transformed P-values.

### Multi-trait Colocalization of CK-associated Loci

We applied Hypothesis Prioritisation for multi-trait Colocalization (HyPrColoc)^66^, a deterministic Bayesian framework designed to identify shared causal variants across multiple GWAS summary statistics. For each locus, we considered a ±500 kb window centered on the lead variant identified in the multi-ancestry meta-analysis (including BBJ), using effect estimates from the European-specific meta-analysis. Allelic orientations were harmonized across traits to ensure consistency. HyPrColoc was executed with default parameters, incorporating effect sizes and their standard errors. Colocalization was considered significant when the posterior probability exceeded 0.8.

## Statements

### Contribution

Conceptualization: SSZ, APM, JAL, GC; Data curation: GC, SSZ, JBL; Methodology: APM, JAL, GC, SSZ, WL; Formal analysis: GC, WL (pQTL MR); Visualization: GC; Supervision: APM, JAL, HC; Writing – original draft: GC; Writing – review & editing: all authors. These authors contributed equally: APM, JAL. Correspondence and requests for materials should be addressed to APM or JAL. All authors approved the final manuscript.

## Data Availability

All data produced in the present study are available upon reasonable request to the authors.

## Acknowledgement

We thank the participants and investigators of the BioBank Japan Project, the Million Veteran Program, the Qatar Genome Program, and the Vanderbilt University Medical Center for their invaluable contributions to this study. This study has been delivered through the National Institute for Health and Care Research (NIHR) Manchester Biomedical Research Centre (NIHR203308). The views expressed are those of the author(s) and not necessarily those of the NIHR or the Department of Health and Social Care.

