## Supplemental Material for "Multi-ancestry Genome-wide Association Study of Creatine Kinase Highlights the Genetic Basis of Muscle Damage"

### Supplementary Note. Extended Description of CK-Associated Genetic Loci

Given the large number of loci identified and the constraints of the main text, we provide here extended descriptions of loci that were only briefly mentioned or omitted due to limited supporting evidence. However, due to pleiotropic effects and the substantial functional overlap among pathways, definitive classification is not possible. The categorization and discussion presented here are informed by existing literature and are intended to offer additional context. It is noted that these loci may not always exert their effects through the nearest mapped genes in the absence of supporting evidence such as colocalization, and alternative mechanisms may underlie their associations.

1. **Muscle Structural Integrity & T-Tubule Architecture**

rs11693851(SPEGNB, upstream): The variant acts as an eQTL in muscle tissue for both SPEG and ASIC4. SPEG encodes a myosin light chain kinase–related protein essential for cytoskeletal integrity, and loss-of-function mutations have been associated with congenital myopathy syndromes^1^.

rs12787021 (CORO1B, 3’ UTR): The gene encoding an actin-binding protein involved in cellular motility—functions as a muscle eQTL for CARNS1. In mammals, CARNS1 (carnosine synthase) supports Ca²⁺ handling and excitation–contraction coupling in heart^2^.

rs11081948 (FHOD3, intronic): The gene is a formin involved in actin dynamics and sarcomere assembly, has been linked to splicing alterations^3^. A splicing-disruptive FHOD3 variant has been associated with severe left-ventricular noncompaction, and FHOD3 dysregulation via RBM20-mediated splicing.^3,4^

rs9727721 (OBSCN, intronic): OBSCN encodes a giant sarcomeric protein essential for maintaining myofibrillar integrity and undergoes extensive alternative splicing during muscle development^5^. Variants in OBSCN has also been associated with lean mass, serum creatinine levels, and susceptibility to rhabdomyolysis^6^.

rs11698868 (MYH7B, intronic): MYH7B encodes both a slow-twitch myosin heavy chain protein that with non-productive Splicing and regulatory non-coding RNAs^7^. In cardiac tissue, alternative splicing of MYH7B produces a long non-coding RNA that modulates myosin isoform composition^7,8^.

rs6089067 (TPX2, intronic): The lead variant maps to a muscle-specific eQTL for COX4I2, MYLK2, and DEFB124. Although TPX2 itself has no established role in muscle, MYLK2 encodes a kinase involved in muscle contraction and has been implicated in cardiomyopathy^9^.

rs11073663 (SEC11A, upstream): This locus is associated with muscle expression of NMB, ALPK3, and WDR73. Colocalization implicates ALPK3 expression in the regulation of CK levels. ALPK3, an M-band kinase and functions as a critical scaffold required for cardiomyocyte structure; its disruption has been linked to cardiomyopathy^10^.

CAV3, CAVIN4 (MURC), and BIN1 coordinate transverse tubule formation—membrane invaginations critical for excitation–contraction coupling in muscle^11^. By directing membrane remodeling during myogenesis, these proteins maintain T-tubule architecture; their dysregulation disrupts this structure and contributes to muscle pathology^11^. Three loci identified in the CK GWAS: rs7601283 (BIN1 and CYP27C1, intergenic): BIN1, a BAR-domain–containing protein, generates ring-like membrane scaffolds that serve as nucleation points for T-tubule formation^12^. rs1236813 (CAVIN4, intronic): Cavin4 is recruited to developing T-tubules and helps remodel caveolar component after BIN1 forms membrane scaffolds that nucleate T-tubules^11^. rs237870 (CAV3, intronic): CAV3, a muscle-specific caveolar protein, supports membrane curvature and localizes to nascent T-tubules^13^.

rs2048677 (KCNJ2 and CASC17, intergenic): This variant is annotated as an eQTL for both KCNJ2 and KCNJ2-AS1 in skeletal muscle. KCNJ2 encodes an inward-rectifier potassium channel, an integral membrane protein implicated in maintaining resting membrane potential. KCNJ2 is linked to Andersen-Tawil syndrome, a disorder that could manifest with episodic muscle weakness and elevated serum CK levels^14^. Another two less relavant ion channel proteins identified in the CK GWAS are rs7921994 (KCNMA1, intronic); The gene encodes the large-conductance calcium- and voltage-activated potassium channel, a transmembrane protein characterized by high potassium selectivity and conductance^15^; rs76752100 (CNNM2, intronic); It encodes a divalent metal cation transporter involved in magnesium homeostasis^16^.

2. **Myogenesis, Splicing Control & Nuclear Architecture**

rs2036291 (MBNL1, intronic): MBNL1is a master regulator of alternative splicing implicated in the pathogenesis of myotonic dystrophy^17^. In this disorder, characterized by myotonia and progressive muscle weakness, aberrant MBNL1-mediated splicing is central, despite frequently normal or mildly elevated serum CK levels^17,18^.

rs3790454 (MEF2D, intronic): The variant overlaps the muscle-specific eQTL of MEF2D. MEF2D undergoes tissue-restricted splicing to generate isoforms critical for muscle differentiation and development^19^.

rs73167046 (MRTFA, intronic): MRTFA is a multifaceted protein that serves as a transcriptional coactivator. It plays a central role in smooth and skeletal muscle differentiation, hematopoiesis, and fibrogenic pathways^20^. Recent evidence implicates its role in regulating PAX7 expression during muscle regeneration and in modulating TAZ expression through a TGF-β1 and p38 MAPK-dependent, Smad3-independent mechanism^20,21^.

rs9894648 (NF1, intronic): NF1 is associated with both Neurofibromatosis type 1 and familial spinal neurofibromatosis in ClinVar, maps to a muscle-specific eQTL, suggesting it modulates NF1 expression in skeletal muscle. Given NF1's essential role in maintaining muscle progenitor function, metabolism, and proteostasis, variation at this site may contribute to elevated CK levels via impaired muscle maintenance or subclinical myopathy^22,23^.

rs1206825 (EYA2, intronic): EYA2, in complex with Six1, promotes adaptive muscle growth by activating mTOR transcription, driving physiological cardiac hypertrophy and preserving mitochondrial integrity^24^.

rs9393602 (RIPOR2, intronic): RIPOR2 is a gene essential for skeletal muscle differentiation and myoblast fusion through filopodia formation, interaction with HDAC6 and dysferlin, and inhibition of RHOA activity^25^.

rs137963475 (FBXO21 and NOS1, intergenic): This locus colocalizes with FBXO21 expression in muscle. FBXO21, an E3 ubiquitin ligase regulated by FoxO3, promotes muscle atrophy, and its inhibition has been shown to reduce proteolysis in denervated muscle^26,27^.

rs1955512 (AKAP6, intronic): The variant is associated with HDL cholesterol levels. AKAP6 is a key scaffold of the nuclear-envelope MTOC in muscle, linking nesprin-1α to centrosomal proteins (AKAP9/PCNT) downstream of myogenin^28,29^.

rs71475991 (NUP160, intronic): The locus colocalizes with muscle eQTLs for C1QTNF4 and MTCH2. Functionally, C1QTNF4 inhibits vascular smooth muscle cell proliferation and migration^30^, while MTCH2 regulates energy homeostasis; its loss in muscle enhances energy expenditure and confers protection against diet-induced obesity^31^.

rs72842207 (BAG3, intronic): The gene which encodes a key protein in skeletal and cardiac muscle. BAG3 maintains structural integrity by regulating autophagy and clearing damaged sarcomeric proteins^32^. Mutations in BAG3 impair these functions, leading to muscle weakness, cardiomyopathy, and neuromuscular disorders such as myofibrillar myopathy^32-34^. Another lead variant, rs332242, of DAPK2 and CIAO2A, intergenic: DAPK2, a calcium/calmodulin-regulated kinase, promotes autophagy by inhibiting mTORC1 and induces apoptosis through its catalytic activity^35,36^.

3. **TGF-β Signaling**

rs6507878 (SMAD7, intronic): SMAD7 is an intracellular inhibitor of TGF-β signaling, crucial for muscle development, cancer risk, and cardiac remodeling. It promotes myogenesis by activating muscle-specific genes^37^. Loss of SMAD7 leads to impaired muscle growth due to unchecked SMAD2/3 activity^37,38^. As a feedback regulator, it restrains SMAD3 signaling, but this control can be suppressed by factors like KLF12, which enhances fibrosis through SMAD3 activation^39,40^.

rs12858234 (KLF12 and LINC00402, intergenic): KLF12 is a Krüppel-like transcriptional repressor involved in development and disease. Recent evidence indicates KLF12 is upregulated in cardiac fibroblasts and aggravates Ang II–induced remodeling by repressing SMAD7 and enhancing TGF-β/SMAD3 signaling in mouse^40^.

rs1153287 (BACH1, intronic): BACH1 is a transcription factor that represses oxidative stress response genes and is linked to disorders like Fanconi anemia, β-thalassemia, and pulmonary fibrosis. It aids muscle regeneration by promoting myoblast proliferation and differentiation through Smad2/3 and FoxO1 repression^41^.

4. **Adrenergic, Insulin/IGF, and Mitochondrial Metabolism**

rs1042713 (ADRB2, exonic): Encodes the beta-2-adrenergic receptor, which responds to catecholamines like epinephrine and regulates cardiovascular, respiratory, and metabolic functions. Variants are linked to asthma, obesity, type 2 diabetes, and may affect muscle performance by influencing oxygen delivery and aerobic capacity^42^.

rs1801690 (APOH, exonic): The APOH locus colocalizes with liver enzyme levels (AST and ALT). It encodes apolipoprotein H, which is involved in lipoprotein metabolism, coagulation, and immune functions, including autoantibody production in antiphospholipid syndrome^43,44^.

rs4805881 (PEPD, intronic): An eQTL in muscle, associated with various traits in GWAS. PEPD encodes a dipeptidase critical for proline recycling and collagen metabolism^45^. Mutations lead to prolidase deficiency, affecting collagen turnover and immune function^45,46^.

rs7182976 (IGF1R, intronic): Encodes the IGF-1 receptor, a key mediator of anabolic and survival signals in muscle via PI3K/Akt and MAPK/ERK pathways^47^. Essential for muscle growth, satellite cell proliferation, differentiation, and protection against atrophy. Knockout models show severe muscle loss despite normal glucose tolerance^48^.

rs2972147 (LOC646736 and MIR5702, intergenic): Maps to an IRS1 eQTL in adipose tissue. IRS1 is a critical adaptor for insulin and IGF-1 receptors, activating PI3K/Akt and AMPK pathways in muscle to regulate glucose uptake, protein synthesis, and growth. Disruption impairs muscle development and insulin sensitivity^49^.

Four additional loci were linked to muscle metabolism: rs38042 (LNPEP, intronic), rs7652475 (XYLB and ACVR2B-AS1, intergenic), rs11837319 (ATP5F1B andPTGES3, intergenic), and rs78660602 (PIK3R1 and LINC02198, intergenic). These genes are involved in key metabolic processes—LNPEP in peptide processing, XYLB in sugar metabolism, ATP5F1B in mitochondrial ATP synthesis, and PIK3R1 in insulin signaling. Notably, PIK3R1 and ATP5F1B are directly implicated in muscle energy homeostasis and function^50,51^.

### Supplementary Figure 1. File- and meta-level quality control by EasyQC

(A) Median standard error versus sqrt(N) across cohorts with varying sample sizes. (B) Genomic inflation factor across cohorts with different sample sizes. (C) P–Z plots comparing reported p-values and those derived from Z-scores for each cohort.

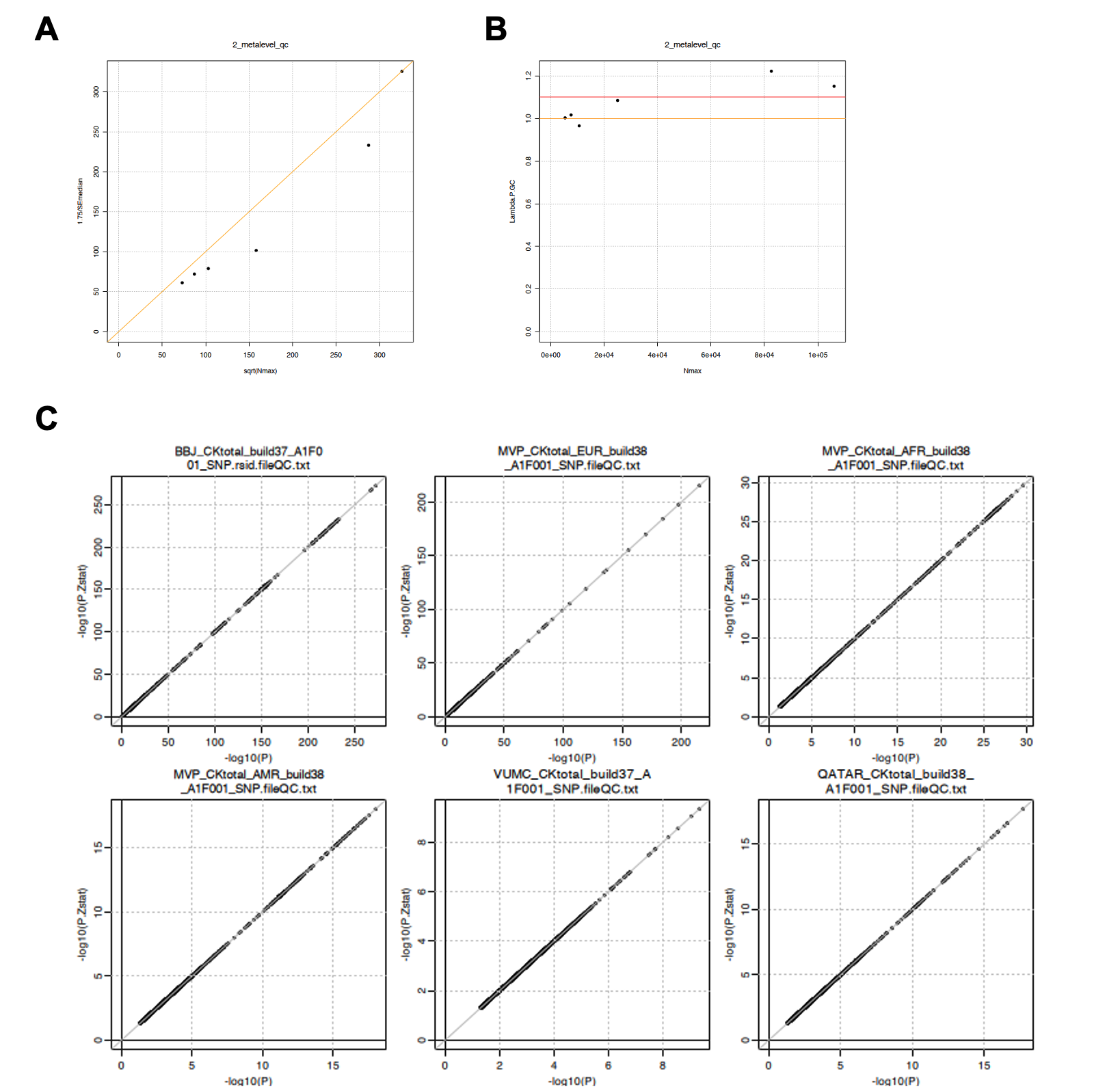

### Supplementary Figure 2. Allele frequency quality control

Minor allele frequencies (MAF) from cohort summary statistics are compared with ancestry-matched 1000 Genomes reference MAF for MVP-EUR, MVP-AMR, MVP-AFR, VUMC, and BBJ. The QGP cohort was compared to 1000 Genomes European reference. The red dashed line indicates perfect concordance.

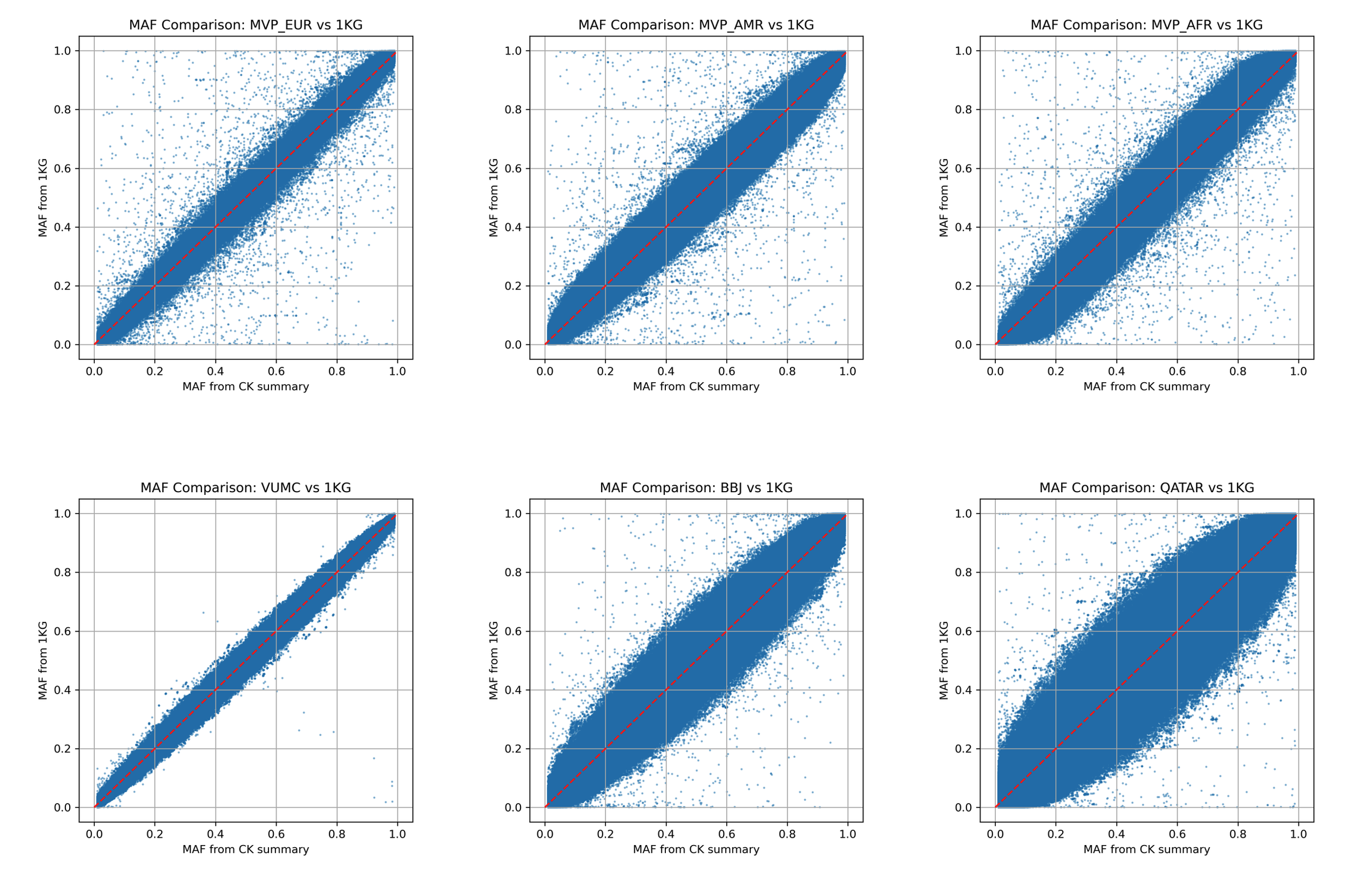

### Supplementary Figure 3. Pairwise comparison of significant SNP effect sizes

Scatter plots show correlations of genome-wide significant SNP effect sizes between cohorts before meta-analysis. Points represent SNPs with 95% confidence intervals. Dashed lines mark zero effect; solid lines show regression slopes with shaded 95% confidence intervals. Pearson’s r and p-values are shown.

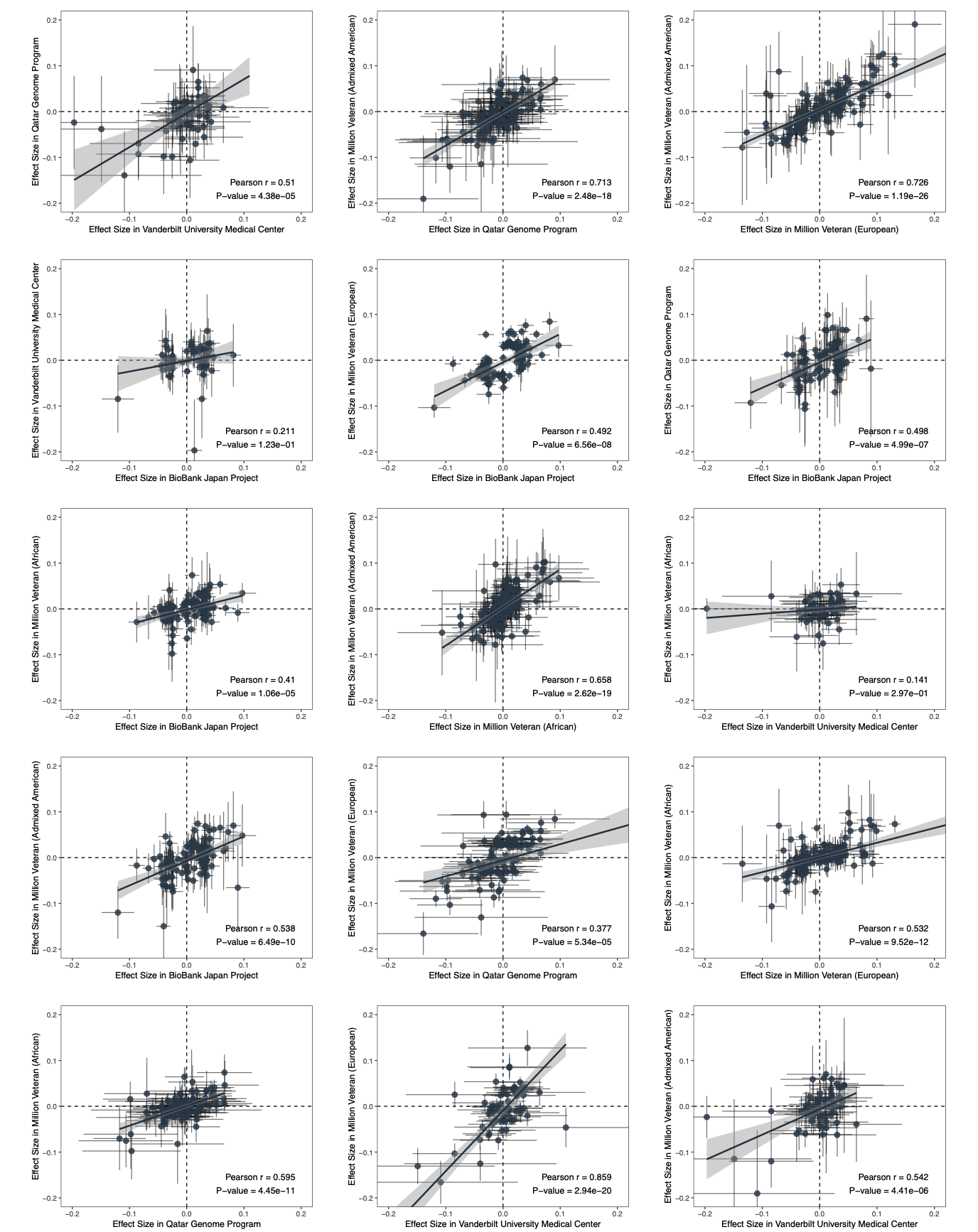

### Supplementary Figure 4. Manhattan plots of cohorts and meta-analyses

Manhattan plots of GWAS results for MVP European, MVP African, MVP Admixed American, BBJ, VUMC, QGP, and standard error–weighted meta-analyses (European and multi-ancestry). The x-axis shows chromosomal positions, and the y-axis shows –log₁₀(p). Red and blue lines denote genome-wide (p = 5 × 10⁻⁸) and suggestive (p = 1 × 10⁻⁶) significance thresholds, respectively.

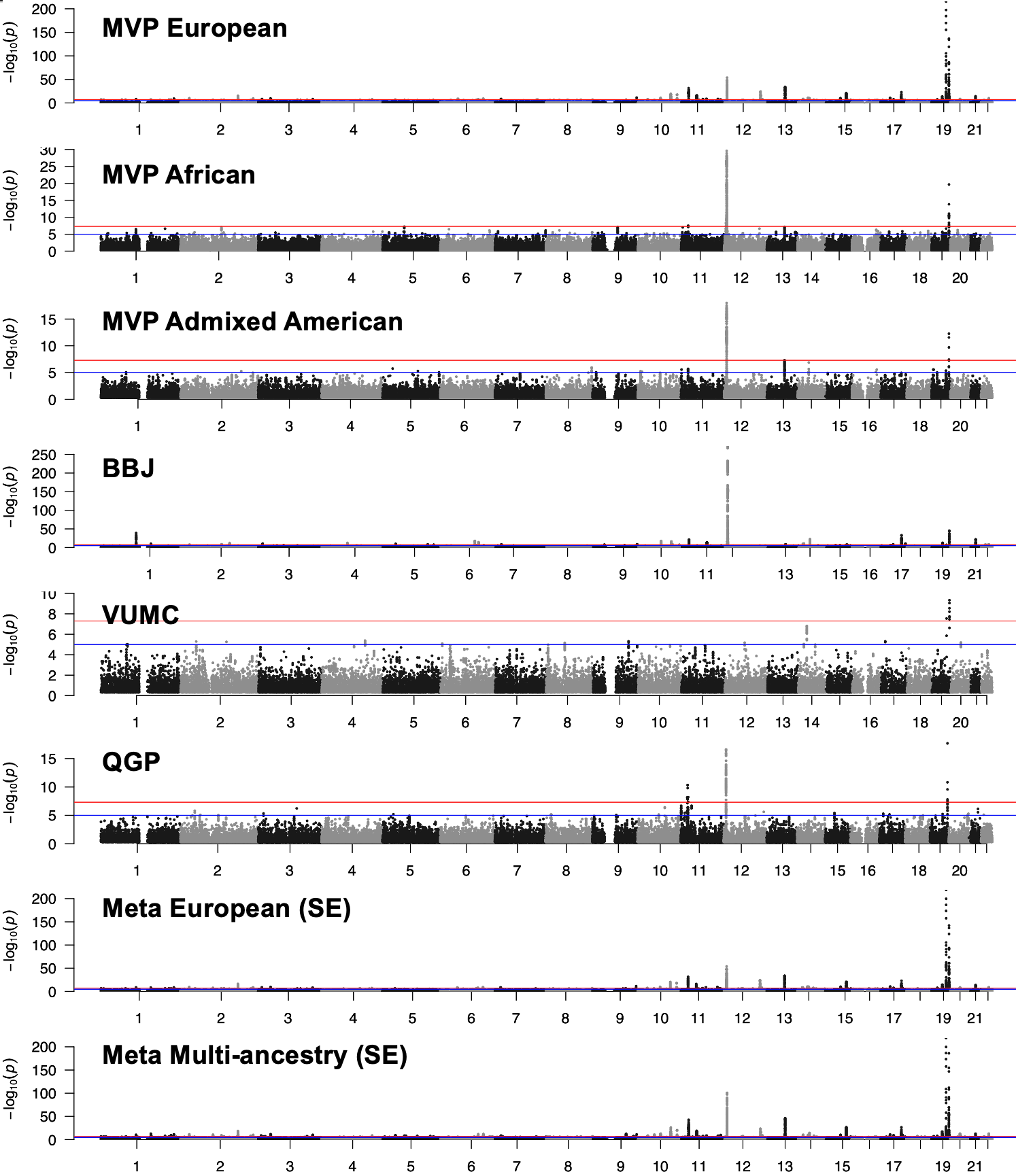

### Supplementary Figure 5. Mendelian randomization plots

Scatter plots show SNP effects for exposures and outcomes with five MR methods: inverse variance weighted, MR Egger, simple mode, weighted median, and weighted mode. Points represent SNP effect estimates, and lines indicate confidence intervals. (A) CK on AST; (B) AST on CK; (C) Hand grip strength on CK; (D) Arm fat-free mass on CK.

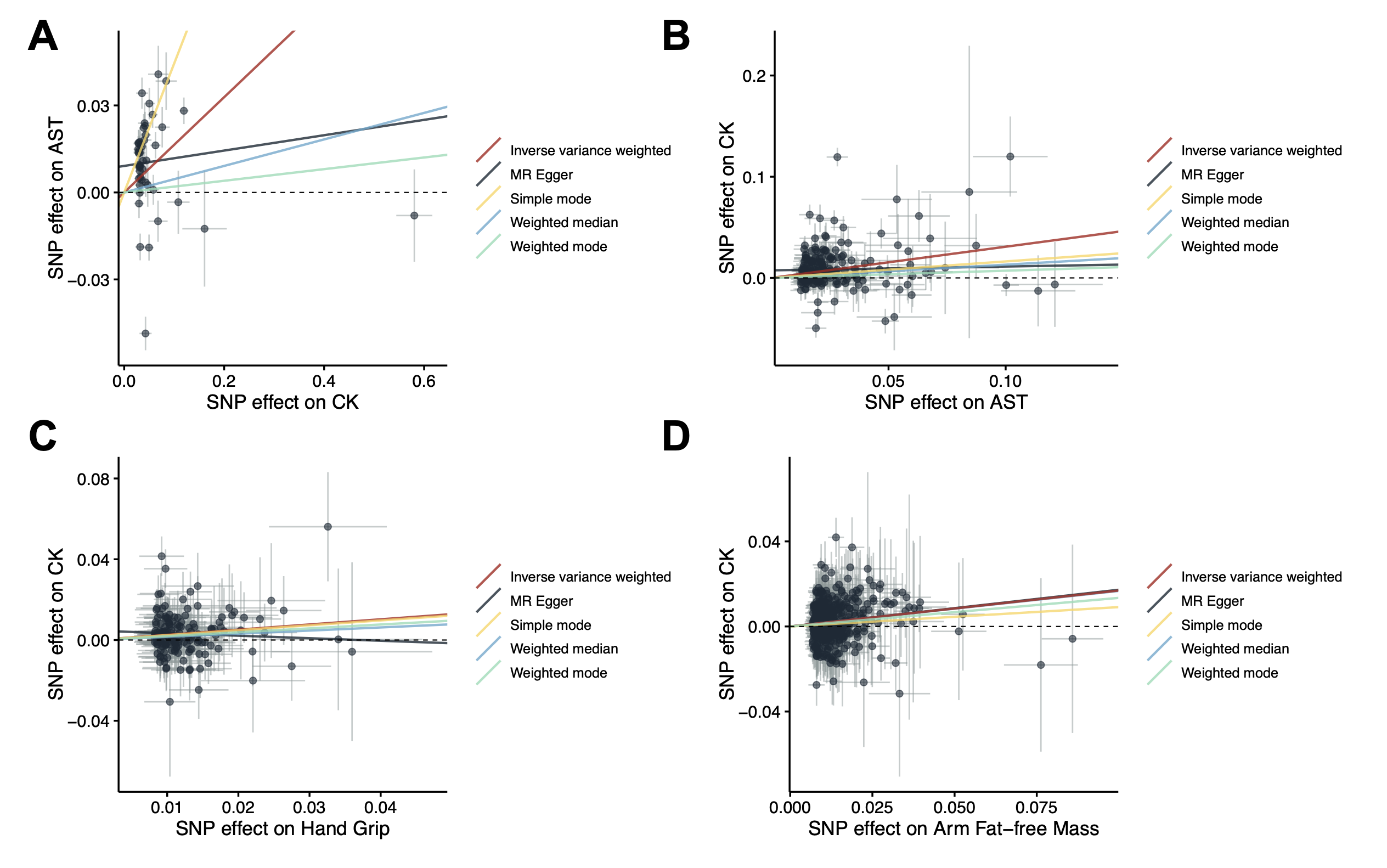

### Supplementary Figure 6. Heatmap of standardized z-scores across traits

Colors indicate effect direction and magnitude (red: consistent with CK, blue: opposite). Rows are loci grouped by trait-based clustering.

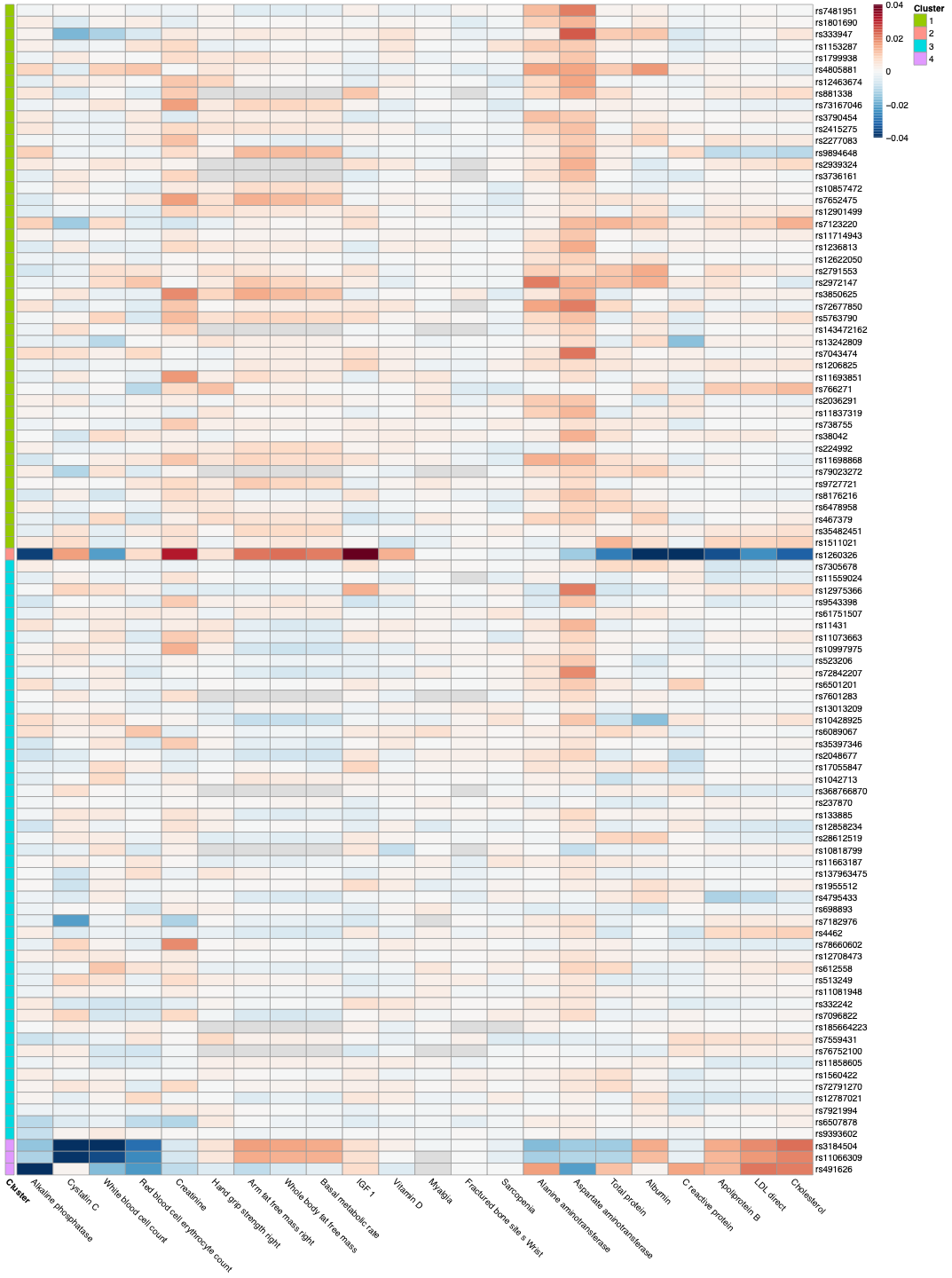
